# Comparison of nasopharyngeal swab and oral fluid collected by ORACOL – Saliva Collection System® for molecular detection of SARS-CoV-2

**DOI:** 10.1101/2024.10.15.24315521

**Authors:** Cyril Barbezange, Veronik Hutse, Inge Roukaerts, Isabelle Thomas, Els Tobback, Steven Callens, Marie-Angélique De Scheerder, Elizaveta Padalko

**Affiliations:** National Influenza Centre, Sciensano, Brussels, Belgium; National Reference Centre for Measles-Mumps-Rubella, Sciensano, Brussels, Belgium; Department of General Internal Medicine, Ghent University Hospital, Ghent, Belgium; Department of Medical Microbiology, Ghent University Hospital, Ghent, Belgium

**Keywords:** SARS-CoV-2, molecular detection, collection device, Oracol, diagnosis COVID-19

## Abstract

The performance of Oracol collection device (Oracol) for oral fluid sampling for SARS-CoV-2 molecular detection was compared to nasopharyngeal swabs (NPS). Samples (n = 128) were collected from symptomatic and asymptomatic adults after PCR confirmed COVID-19 on NPS. All samples were tested by 3 real-time PCR’s (RT-qPCR) for E, N and RdPR genes. Accuracy of Oracol compared to NPS was 38.28 % for E-gene, 47.66 % for RdRp-gene and 83.59 % for N-gene. A clear correlation between the N-gene Ct values on NPS and on Oracols was observed (p < 0.001). The sensitivity of the Oracol compared to the NPS increased in participants with an N-gene Ct value <30 on NPS (89.08%,) reaching 93.2 % with a N-gene Ct value <25. Oracol can be considered an alternative to NPS to detect SARS-CoV-2, especially in populations where NPS is not well tolerated.

## Background

At the end of 2019, a new human coronavirus, now known as severe acute respiratory syndrome coronavirus 2 (SARS-CoV-2), was first reported and is a cause of a currently ongoing pandemic of coronavirus disease 2019 (COVID-19) with the clinical presentations ranging from no to limited symptoms to patients with severe potentially life-threatening pneumonia requiring hospitalization [1].

At the time of writing, the WHO reported more than 440 million infections and a death toll of more than 5.5 million since the start of the pandemic. The rise of the virus and the economic and social impact on the whole world caused a surge of research and the rapid development of several effective vaccines. To date more than 10 billion vaccine doses have been administered worldwide, helping in quenching the impact of the pandemic.

Hygienic measures across the world rely heavily on effective testing. Testing is first performs to confirm an infection in a symptomatic patient to decide on the therapeutic path. Second, testing might be performed to identify infectious individuals in a population, who are then isolated to prevent further spread. Ending the pandemic involves the accurate application of diagnostic testing in high volumes and the rapid use of the results to help implement the appropriate therapy and prevent further spread.

In January 2020 the genome of the virus was published, and within two weeks the first diagnostic PCR tests were available [2]. A PCR for RdRp, E, N or S genes was recommended and is performed on nasopharyngeal swabs [3]. Based on established diagnostics for other respiratory infections, the nasopharyngeal swab was initially adopted as the preferred sampling technique. However, numerous reports have shown that saliva can serve as an alternative upper respiratory tract specimen type for SARS-COV-2 detection [4–8].

Nasopharyngeal sampling requires a swab being inserted into the back of the nose, which can cause irritation that could promote sneezing and coughing. Thus, the non-invasive collection of saliva would be safer, as it protects healthcare workers from being inadvertently exposed to potentially infectious aerosols. This less irritating way of sampling is also especially interesting for people that require frequent testing or for children.

In this study, we evaluated the performance of sampling by Oracol collection device (Malvern Medical Development, UK) (Oracol) compared to nasopharyngeal sampling (NPS) to detect SARS-CoV-2 virus for the COVID-19 diagnosis by PCR. Oracol sampling has been routinely used for diagnostic purposes for the detection of measles and mumps viruses by RT-PCR, as well as for the detection of antibodies against measles, mumps, rubella viruses [9]. Oracol collection device was specifically developed for the collection of oral fluid enriched in gingival crevicular fluid and thus differs from pure saliva obtained from spitting and from saliva mixed with sputum [10]. It is a painless, non-invasive sampling procedure and could thus represent an interesting alternative to nasopharyngeal swab that causes more discomfort, especially for children. Both NPS and Oracol samples were tested with a PCR for the SARS-CoV-2 E-gene, N-gene and the RdRp-gene.

## Methods

### Samples and participants

Study samples were collected from the participants of the simultaneously ongoing CAMOSTAT study [11] which included repetitive visits on day 5 and day 10 after initial diagnosis of COVID-19 infection by a polymerase chain reaction (PCR) on a nasopharyngeal swab (NPS). The Oracol study participants were asked if they could provide, in addition to the NPS already included in the CAMOSTAT study, a saliva sample collected by ORACOL – Saliva Collection System® during the same visit. Ethical approval from all of the appropriate institutional review boards was obtained. The ethics review committee of Ghent University Hospital Belgium, approved the study with a reference number B670202000996.

NPS samples were collected by a nurse using eSwab Mini Tip FloQ swab®, Copan (Brescia, Italy), according to the manufacturer’s instructions by gently inserting the swab into the nostril and pushing it deeper into the pharynx with the patient’s head slightly tilted backwards.

Saliva samples were collected by the patient himself using ORACOL – Saliva Collection System® (Malvern Medical Developments, Worchester, United Kingdom) by placing the collection device into the mouth with consequent continuous rotation between cheek and upper teeth during minimal 1, maximal 2 minute(s).

After collection, the swabs were placed in the provided plastic tubes and transported to the Laboratory of Medical Microbiology, Ghent University Hospital, where the saliva samples were processed by centrifugation at 3000 g for 10 minutes in order to collect the patient’s saliva absorbed by the sponge swab. All samples were stored at −80°C temperature prior to dry-ice transport to Sciensano for molecular testing.

### Molecular assays

All collected NPS and saliva samples were tested at Sciensano by 3 real-time PCR methods accordingly to previously published procedures targeting separately the E-gene, N-gene (US CDC N1 protocol) and RdRp-gene (Institut Pasteur protocol) of SARS-CoV-2 [3]. PCR’s were performed after nucleic acid extraction using Nuclisens® easymag® reagents on an EMAG® instrument (Biomérieux).

### Evaluation of results

Data processing and statistical analysis was performed in R program version 4.1.1 and graphs were created using GraphPad Prism version 9.0.0.

## Results

During the course of the CAMOSTAT study, 134 participants were enrolled in this sub-study to evaluate the performance of Oracol sampling to detect SARS-CoV-2 virus in oral fluid samples. Participants for the Oracol sub-study were enrolled among the CAMOSTAT outpatients (not hospitalized) who tested positive for SARS-CoV-2 and who received the placebo treatment. They were then contacted to take part in the Oracol sub-study. At enrollment in the sub-study, a nasopharyngeal swab and an Oracol swab were collected by and under the supervision of a nurse, respectively.

All samples were tested with three RT-qPCRs targeting different genes of the virus. Among these 134 participants, only 128 tested positive by RT-qPCR for SARS-CoV-2 (whatever the target) in the nasopharyngeal swab taken on the same day as the Oracol and were thus retained in the analysis to evaluate the performance of Oracol compared to nasopharyngeal swab.

The sub-study population was equally distributed between males and females (64 participants in each group, Table 1). Female participants were on average younger than males (Wilcoxon signed-rank test, p = 0.007), with a clear over representation of females aged below 30 (Fisher exact test, p = 0.013). There was suggestive evidence of a difference between gender in the proportion of symptomatic patients (Fisher exact test, p = 0.054), but not in the number of days after symptom onset. There was strong evidence that a sore throat was more often reported by females (Fisher exact test, p = 002) and suggestive evidence for sneezing and headache (Table 1). There was no difference between age groups in the proportion of symptomatic patients or in the number of days after symptom onset, but there was strong evidence that younger age groups seemed to report sneezing, headache and a sore throat more often than older ones, and that older age groups seemed to report fever more often than younger ones (Table 2).

**Table 1.** Participant characteristics (age and symptomatic status) per gender.

|  |  | gender |  |  | p-value |
| --- | --- | --- | --- | --- | --- |
|  |  | total | male | female |  |
| total |  | 128 | 64 | 64 |  |
| age<sup>a</sup> group | <30 | 54 | 18 | 36 | **0.013<sup>b</sup>** |
|  | 30-40 | 19 | 11 | 8 |  |
|  | 40-50 | 20 | 12 | 8 |  |
|  | 50-60 | 14 | 11 | 3 |  |
|  | 60-70 | 15 | 10 | 5 |  |
|  | 70+ | 6 | 2 | 4 |  |
| age | min | 20 | 20 | 20 | **0.007<sup>c</sup>** |
|  | 1st quartile | 24 | 26 | 22 |  |
|  | median | 37 | 44 | 26.5 |  |
|  | 3rd quartile | 53 | 55 | 42.5 |  |
|  | max | 89 | 75 | 89 |  |
|  | mean | 39.27 | 42.97 | 35.56 |  |
| symptomatic | yes | 117 | 62 | 55 | **0.054<sup>b</sup>** |
|  | no | 11 | 2 | 9 |  |
| onset<sup>d</sup> | min | 0 | 0 | 0 | 0.700<sup>c</sup> |
|  | 1st quartile | 1 | 1 | 2 |  |
|  | median | 4 | 3.5 | 5 |  |
|  | 3rd quartile | 6 | 6 | 6 |  |
|  | max | 14 | 14 | 14 |  |
|  | mean | 4.22 | 4.18 | 4.27 |  |
| symptoms | cough | 65 | 37 | 28 | 0.358<sup>b</sup> |
|  | myalgia | 28 | 15 | 13 | 1.000<sup>b</sup> |
|  | fever | 37 | 17 | 20 | 0.325<sup>b</sup> |
|  | dyspnea | 14 | 5 | 9 | 0.254<sup>b</sup> |
|  | sneezing | 73 | 34 | 39 | **0.087<sup>b</sup>** |
|  | anosmia | 38 | 21 | 17 | 0.874<sup>b</sup> |
|  | dysgeusia | 38 | 21 | 17 | 0.874<sup>b</sup> |
|  | headache | 65 | 29 | 36 | **0.062<sup>b</sup>** |
|  | gastrointestinal | 26 | 11 | 15 | 0.267<sup>b</sup> |
|  | sore throat | 27 | 7 | 20 | **0.002<sup>b</sup>** |

<sup>a</sup> in years
<sup>b</sup> two-sided Fisher exact test
<sup>c</sup> Wilcoxon signer-rank test
<sup>d</sup> among symptomatic (in days after symptom onset)

**Table 2.**
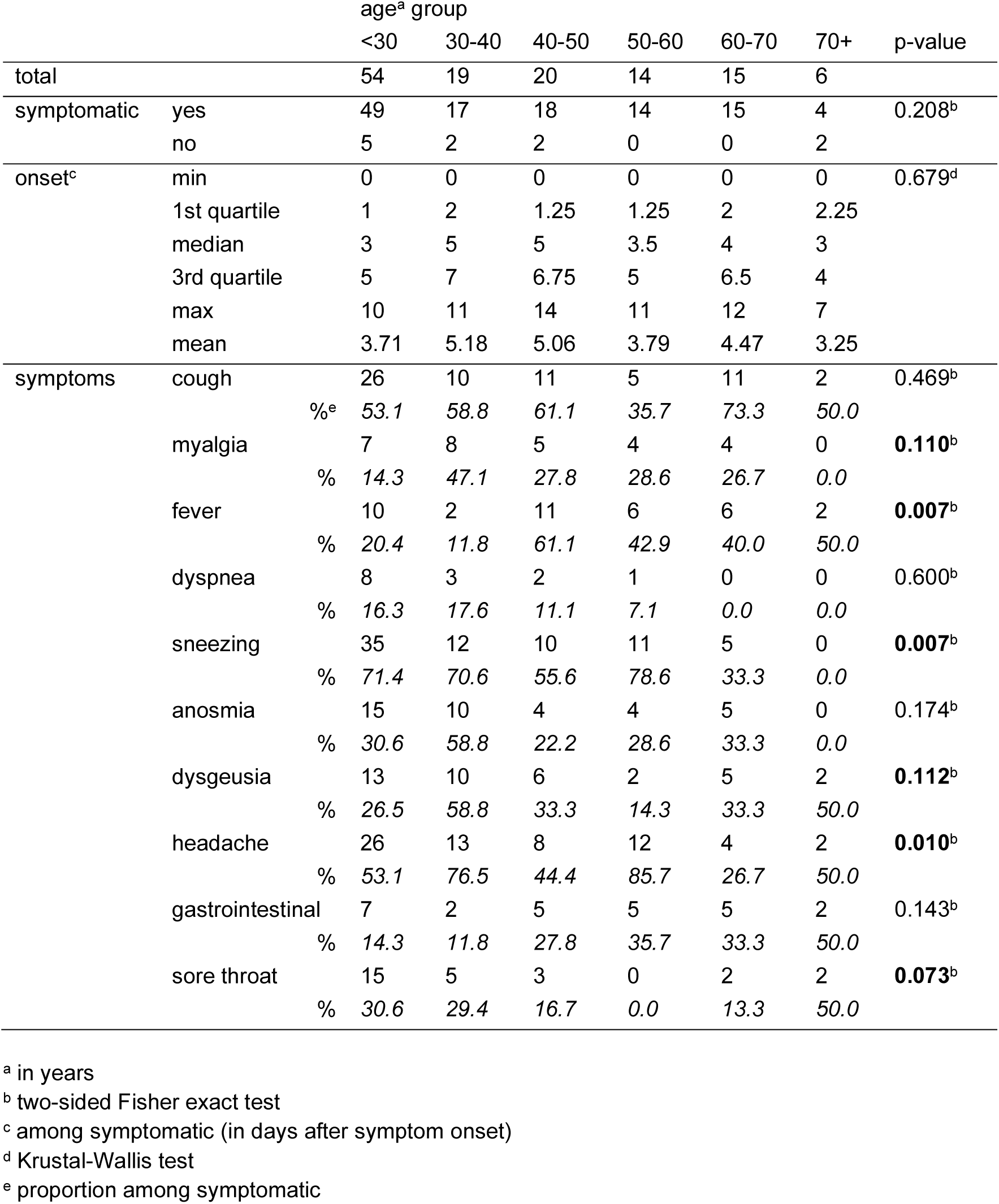
Symptomatic status per age group.

|  |  | age<sup>a</sup> group |  |  |  |  |  | p-value |
| --- | --- | --- | --- | --- | --- | --- | --- | --- |
|  |  | <30 | 30-40 | 40-50 | 50-60 | 60-70 | 70+ |  |
| total |  | 54 | 19 | 20 | 14 | 15 | 6 |  |
| symptomatic | yes | 49 | 17 | 18 | 14 | 15 | 4 | 0.208<sup>b</sup> |
|  | no | 5 | 2 | 2 | 0 | 0 | 2 |  |
| onset<sup>c</sup> | min | 0 | 0 | 0 | 0 | 0 | 0 | 0.679<sup>d</sup> |
|  | 1st quartile | 1 | 2 | 1.25 | 1.25 | 2 | 2.25 |  |
|  | median | 3 | 5 | 5 | 3.5 | 4 | 3 |  |
|  | 3rd quartile | 5 | 7 | 6.75 | 5 | 6.5 | 4 |  |
|  | max | 10 | 11 | 14 | 11 | 12 | 7 |  |
|  | mean | 3.71 | 5.18 | 5.06 | 3.79 | 4.47 | 3.25 |  |
| symptoms | cough | 26 | 10 | 11 | 5 | 11 | 2 | 0.469<sup>b</sup> |
|  | %<sup>e</sup> | 53.1 | 58.8 | 61.1 | 35.7 | 73.3 | 50.0 |  |
|  | myalgia | 7 | 8 | 5 | 4 | 4 | 0 | **0.110<sup>b</sup>** |
|  | % | 14.3 | 47.1 | 27.8 | 28.6 | 26.7 | 0.0 |  |
|  | fever | 10 | 2 | 11 | 6 | 6 | 2 | **0.007<sup>b</sup>** |
|  | % | 20.4 | 11.8 | 61.1 | 42.9 | 40.0 | 50.0 |  |
|  | dyspnea | 8 | 3 | 2 | 1 | 0 | 0 | 0.600<sup>b</sup> |
|  | % | 16.3 | 17.6 | 11.1 | 7.1 | 0.0 | 0.0 |  |
|  | sneezing | 35 | 12 | 10 | 11 | 5 | 0 | **0.007<sup>b</sup>** |
|  | % | 71.4 | 70.6 | 55.6 | 78.6 | 33.3 | 0.0 |  |
|  | anosmia | 15 | 10 | 4 | 4 | 5 | 0 | 0.174<sup>b</sup> |
|  | % | 30.6 | 58.8 | 22.2 | 28.6 | 33.3 | 0.0 |  |
|  | dysgeusia | 13 | 10 | 6 | 2 | 5 | 2 | **0.112<sup>b</sup>** |
|  | % | 26.5 | 58.8 | 33.3 | 14.3 | 33.3 | 50.0 |  |
|  | headache | 26 | 13 | 8 | 12 | 4 | 2 | **0.010<sup>b</sup>** |
|  | % | 53.1 | 76.5 | 44.4 | 85.7 | 26.7 | 50.0 |  |
|  | gastrointestinal | 7 | 2 | 5 | 5 | 5 | 2 | 0.143<sup>b</sup> |
|  | % | 14.3 | 11.8 | 27.8 | 35.7 | 33.3 | 50.0 |  |
|  | sore throat | 15 | 5 | 3 | 0 | 2 | 2 | **0.073<sup>b</sup>** |
|  | % | 30.6 | 29.4 | 16.7 | 0.0 | 13.3 | 50.0 |  |

<sup>a</sup> in years<sup>b</sup> two-sided Fisher exact test<sup>c</sup> among symptomatic (in days after symptom onset)<sup>d</sup> Krustal-Wallis test<sup>e</sup> proportion among symptomatic

All 128 nasopharyngeal swabs were found positive by N-gene RT-qPCR (100 %, CI = 97.09 – 100.00). Only 118 were detected by the E-gene RT-qPCR (sensitivity compared to N-gene = 92.19 %, CI = 86.22 – 95.70) and 121 by the RdRp-gene RT-qPCR (sensitivity compared to N-gene = 95.31 %, CI = 90.15 – 97.83).

When comparing Oracols to nasopharyngeal swabs, the E-gene and RdRp-gene RT-qPCRs showed low accuracy even for samples with a low Ct value (i.e. high viral load) in the nasopharyngeal swab (Figure 1). Accuracy of Oracols compared to nasopharyngeal swabs was only 38.28 % (CI = 29.83 – 47.28) if using the E-gene RT-qPCR, and reached nearly 50 % (47.66%, CI = 38.76 – 56.66) if using the RdRp-gene RT-qPCR. Using the N-gene RT-PCR, more than three quarters of the participants were correctly identified in the Oracol sample (sensitivity compared to nasopharyngeal swab = 83.59 %, CI = 76.22 – 89.01). There was even strong evidence of correlation between the N-gene Ct values obtained for the nasopharyngeal and the Oracol samples (Spearman rank test, rho(126) = 0.625, p < 0.001). The sensitivity of the Oracol compared to the nasopharyngeal swab was increased when considering only participants with an N-gene Ct value <30 in the nasopharyngeal swab (89.08%, CI: 82.2 – 93.5, n = 119) and even reached 93.2 % (CI: 86.63 – 96.67) when considering only participants with an N-gene Ct value <25 (n = 103).

**Figure 1.**
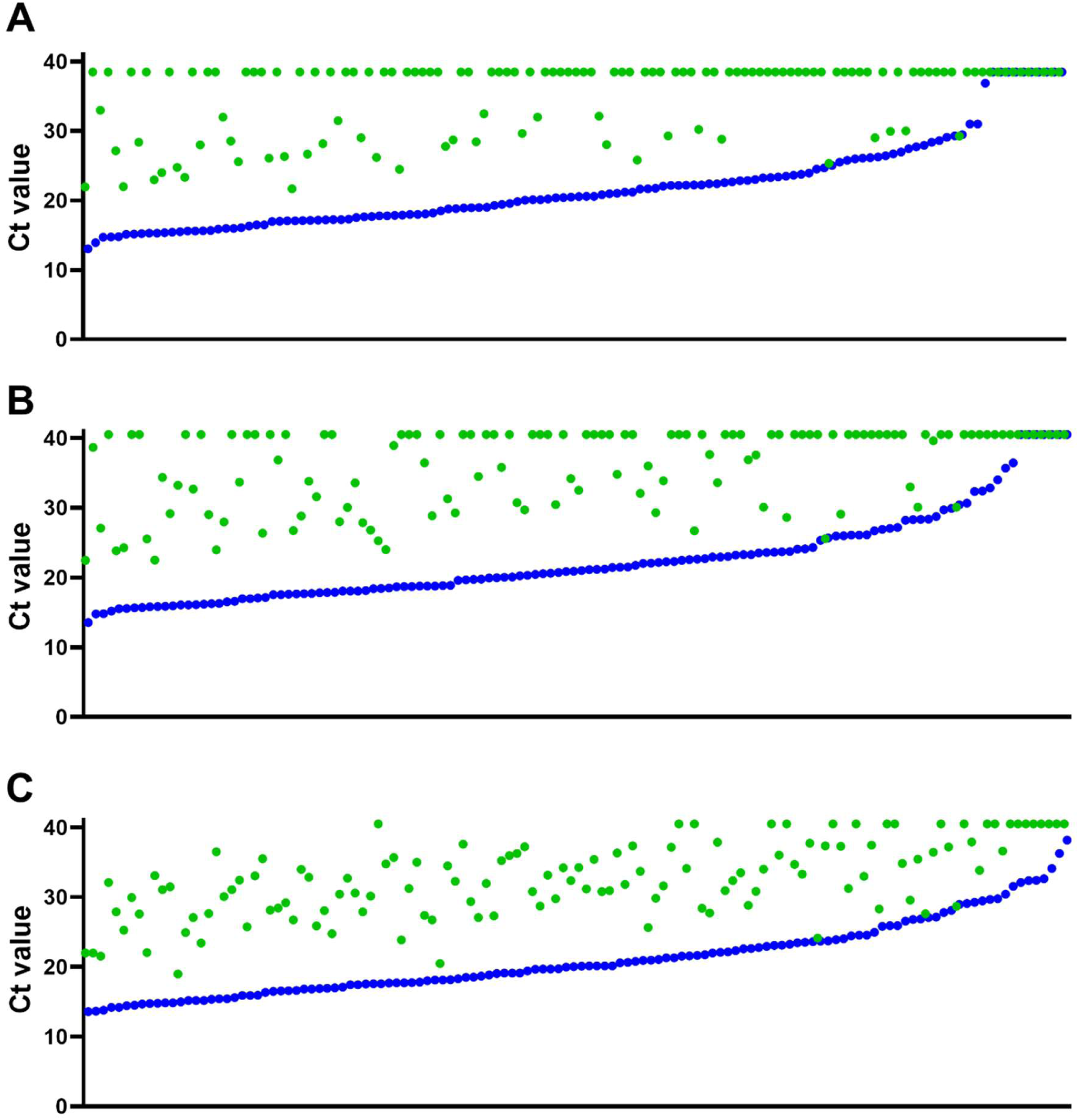
Correlation between Ct values obtained for each target in nasopharyngeal sample (blue) and Oracol (green). A) E-gene RT-qPCR: detection threshold Ct = 38 (not detected were assigned 38.5). B) RdRp-gene RT-qPCR and C) N-gene RT-qPCR: detection threshold Ct = 40 (not detected were assigned 40.5). X-axis: samples were ordered in increasing Ct value in the nasopharyngeal sample.

The association of the N-gene result in Oracol with several univariate explanatory variables was then evaluated (Table 3, Figure 2). The proportion of positivity in Oracol did not significantly differ by gender (chi-square test, x^2^(1, N=128) = 1.42, p = 0.233) – although a Wilcoxon signed-rank test revealed some evidence of a difference between males and females in Ct value obtained (p = 0.040) – nor with age group (chi-square for trend, x^2^(1, N=128) = 1.54, p = 0.215). There was suggestive evidence that the proportion of positivity in Oracol was higher in the group of symptomatic patients compared to the asymptomatic ones (chi-square test, x^2^(1, N=128) = 3.50, p = 0.062). When considering only these symptomatic patients, there was a strong evidence that the proportion of positivity in Oracol differed between symptom-onset groups defined as sampling taking place less than 3 days, between 3 and 5 days, or more than 5 days after onset (chi-square for trend, x^2^(1, N=117) = 9.53, p = 0.002). When considering each symptom separately, there was strong evidence that the proportion of N-gene Oracol positivity was lower in the groups presenting dyspnea (Fisher’s exact test, p = 0.031) or dysgeusia (chi-square test, x^2^(1, N=117) = 6.30, p = 0.012).

**Figure 2.**
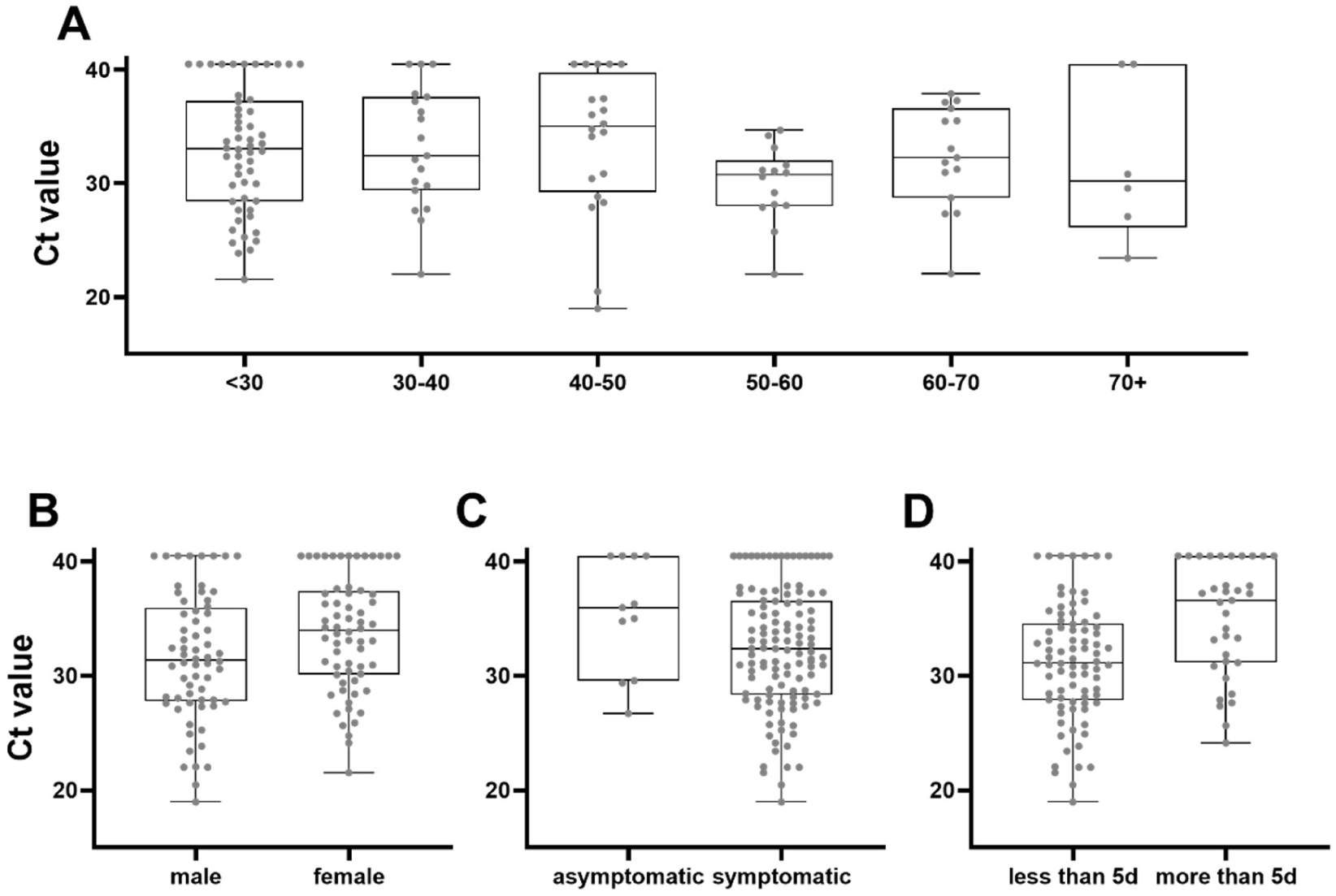
Distribution of N-gene RT-qPCR Ct-values in Oracol visualized per group. A) age group (N=128); B) gender (N=128); C) symptomatic status (N=128); D) days after symptom onset (N=117).

**Table 3:**
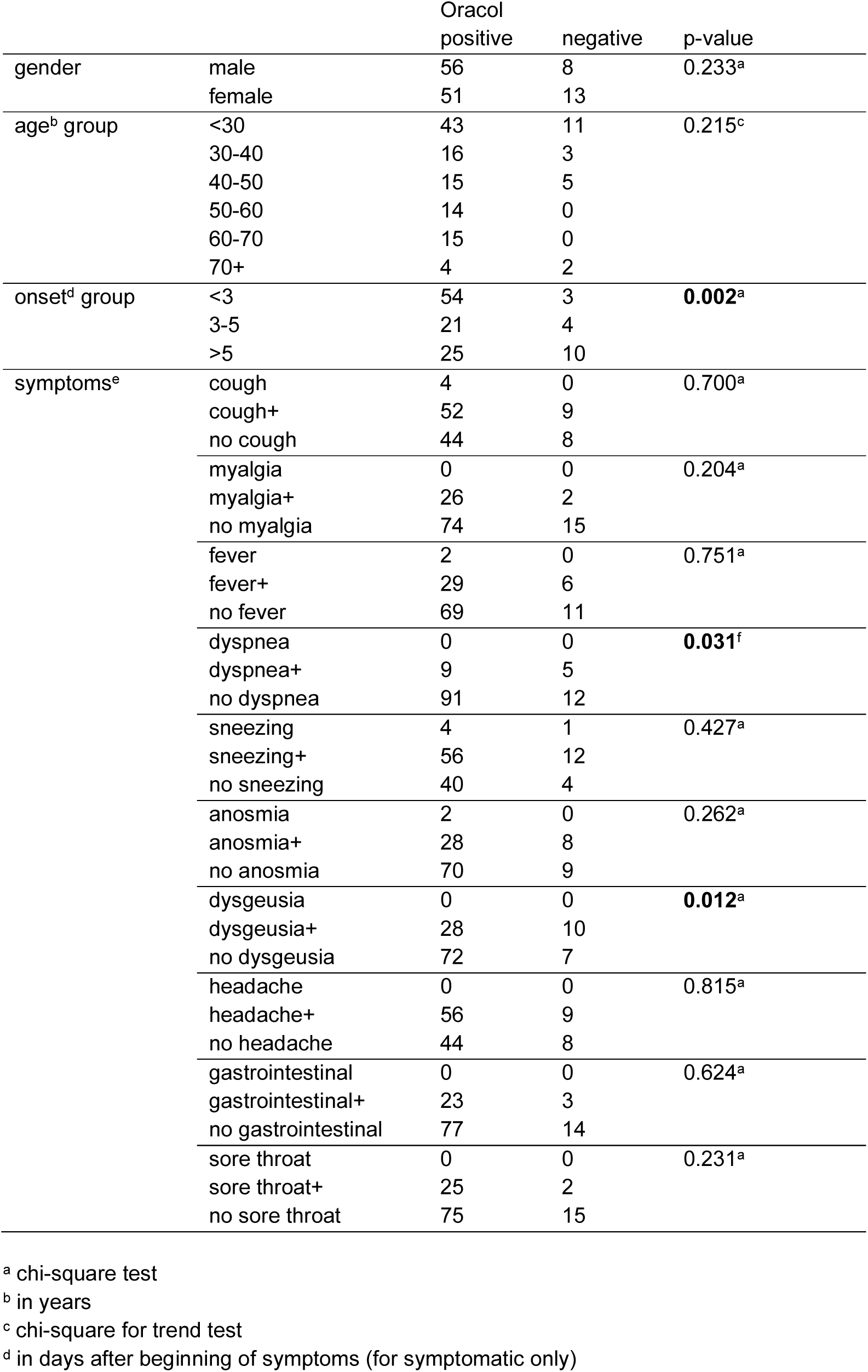
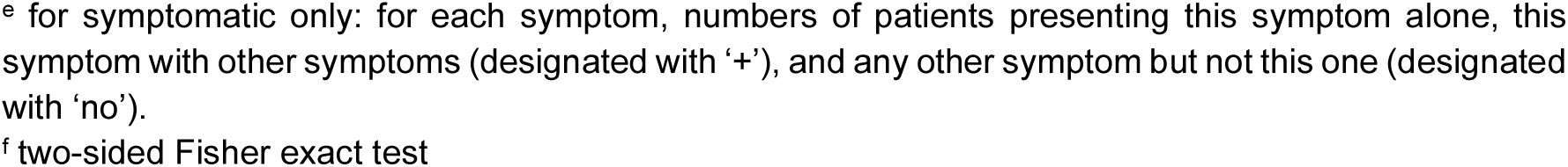
Oracol N-gene positivity status.

|  |  | Oracol<br>positive | negative | p-value |
| --- | --- | --- | --- | --- |
| gender | male | 56 | 8 | 0.233<sup>a</sup> |
|  | female | 51 | 13 |  |
| age<sup>b</sup> group | <30 | 43 | 11 | 0.215<sup>c</sup> |
|  | 30-40 | 16 | 3 |  |
|  | 40-50 | 15 | 5 |  |
|  | 50-60 | 14 | 0 |  |
|  | 60-70 | 15 | 0 |  |
|  | 70+ | 4 | 2 |  |
| onset<sup>d</sup> group | <3 | 54 | 3 | **0.002<sup>a</sup>** |
|  | 3-5 | 21 | 4 |  |
|  | >5 | 25 | 10 |  |
| symptoms<sup>e</sup> | cough | 4 | 0 | 0.700<sup>a</sup> |
|  | cough+ | 52 | 9 |  |
|  | no cough | 44 | 8 |  |
|  | myalgia | 0 | 0 | 0.204<sup>a</sup> |
|  | myalgia+ | 26 | 2 |  |
|  | no myalgia | 74 | 15 |  |
|  | fever | 2 | 0 | 0.751<sup>a</sup> |
|  | fever+ | 29 | 6 |  |
|  | no fever | 69 | 11 |  |
|  | dyspnea | 0 | 0 | **0.031<sup>f</sup>** |
|  | dyspnea+ | 9 | 5 |  |
|  | no dyspnea | 91 | 12 |  |
|  | sneezing | 4 | 1 | 0.427<sup>a</sup> |
|  | sneezing+ | 56 | 12 |  |
|  | no sneezing | 40 | 4 |  |
|  | anosmia | 2 | 0 | 0.262<sup>a</sup> |
|  | anosmia+ | 28 | 8 |  |
|  | no anosmia | 70 | 9 |  |
|  | dysgeusia | 0 | 0 | **0.012<sup>a</sup>** |
|  | dysgeusia+ | 28 | 10 |  |
|  | no dysgeusia | 72 | 7 |  |
|  | headache | 0 | 0 | 0.815<sup>a</sup> |
|  | headache+ | 56 | 9 |  |
|  | no headache | 44 | 8 |  |
|  | gastrointestinal | 0 | 0 | 0.624<sup>a</sup> |
|  | gastrointestinal+ | 23 | 3 |  |
|  | no gastrointestinal | 77 | 14 |  |
|  | sore throat | 0 | 0 | 0.231<sup>a</sup> |
|  | sore throat+ | 25 | 2 |  |
|  | no sore throat | 75 | 15 |  |

<sup>a</sup> chi-square test<sup>b</sup> in years<sup>c</sup> chi-square for trend test<sup>d</sup> in days after beginning of symptoms (for symptomatic only)
<sup>e</sup> for symptomatic only: for each symptom, numbers of patients presenting this symptom alone, this symptom with other symptoms (designated with '+'), and any other symptom but not this one (designated with 'no').
<sup>f</sup> two-sided Fisher exact test

Multivariate logistic regression analyses were then used to identify potential predictors of positivity by N-gene RT-PCR in Oracol samples, taking into account cofounder effects (Table 4, Figure 3). The results of the analysis for the whole study population (N = 128) indicated that, all else being equal, participants with a Ct value below 25 or 30 in the nasopharyngeal swab had higher odds of being detected positive in the Oracol sample than participants with a Ct value equal or above 25 or 30 in the nasopharyngeal swab (adjusted OR (aOR) = 19.77, CI = 6.36 – 70.75, p < 0.001 and aOR = 74.08, CI = 10.85 – 1584.96, p < 0.001, respectively). Among symptomatic participants (N = 117), all else being equal, there was strong evidence that participants with dysgeusia or being sampled more than 5 days after symptom onset had lower odds of being detected positive in the Oracol sample (aOR = 0.24, CI = 0.07 – 0.77, p = 0.019 and aOR = 0.22, CI = 0.06 – 0.69, p = 0.011, respectively), and suggestive evidence for participants with dyspnea (OR = 0.26, CI = 0.06 – 1.14, p = 0.065).

**Figure 3.**
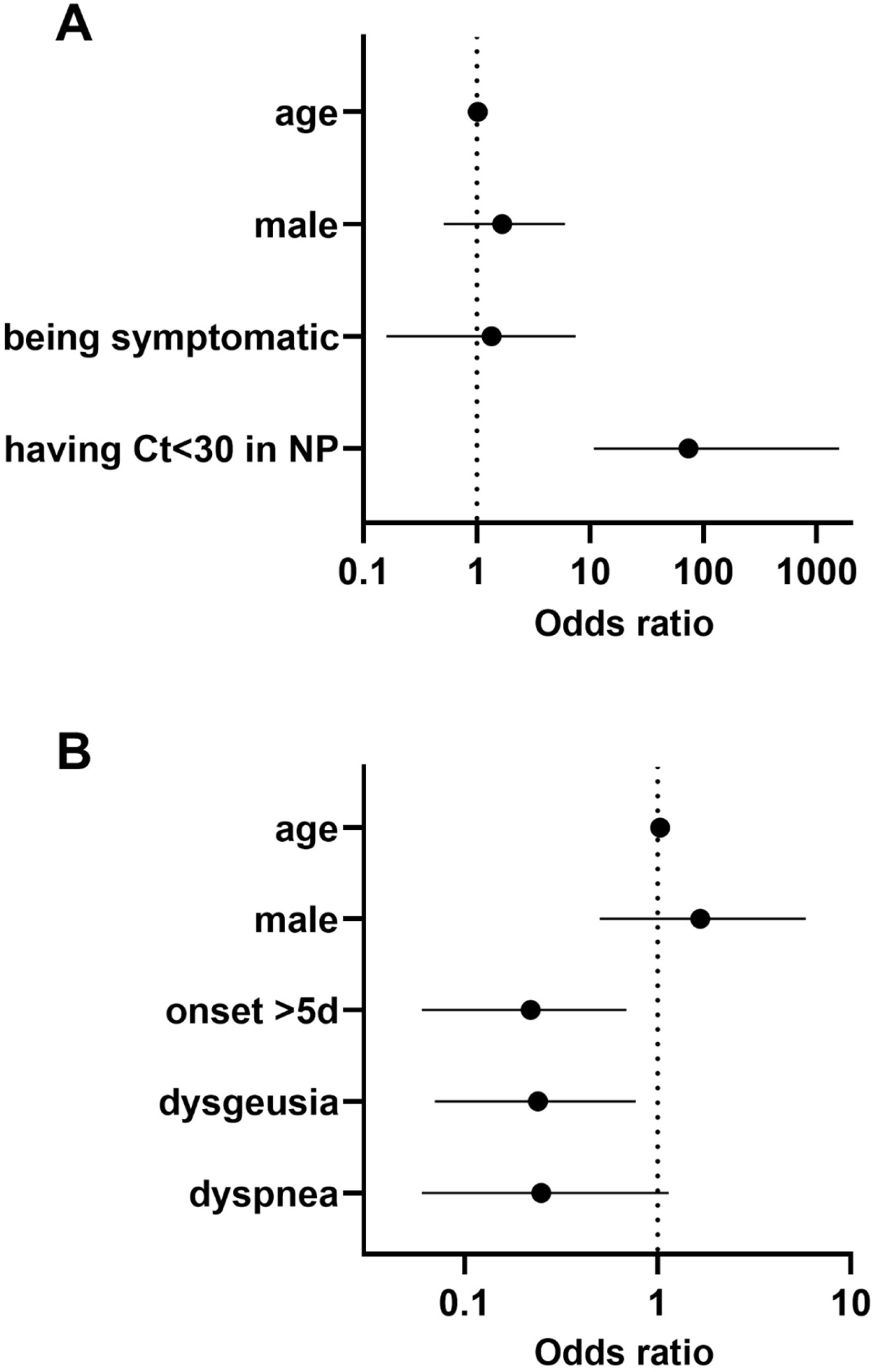
Odds ratios obtained by multivariate logistic regression. A) Model 1 for the whole study population (N = 128): dependent variable = N-gene RT-qPCR positivity status in Oracol sample; explanatory variables = age (continuous variable), gender (binary, male compared to female), symptomatic status (binary, symptomatic compared to asymptomatic), N-gene RT-qPCR Ct-value in matching nasopharyngeal swab (binary, “low”/”high” using a Ct threshold of 30). B) Model 2 for symptomatic participants (N = 117): dependent variable = N-gene RT-qPCR positivity status in Oracol sample; explanatory variables = age (continuous variable), gender (binary, male compared to female), symptom onset status (binary, sampling less compared to more than 5 days after beginning of symptoms), dysgeusia (binary, with compared to without), dyspnea (binary, with compared to without).

**Table 4:** Multiple logistic regression model.

|  | Variable | B | SE | z | p-value | aOR | 95%CI |
| --- | --- | --- | --- | --- | --- | --- | --- |
| Model 1<sup>a</sup> | (Intercept) | -3.469 | 1.539 | -2.254 | 0.024 | 0.03 | 0.00-0.48 |
|  | Age | 0.021 | 0.019 | 1.095 | 0.274 | 1.02 | 0.99-1.06 |
|  | Gender male | 0.520 | 0.617 | 0.842 | 0.400 | 1.68 | 0.51-6.02 |
|  | Symptomatic | 0.298 | 0.945 | 0.315 | 0.753 | 1.35 | 0.16-7.49 |
|  | gpCt30low | 4.305 | 1.169 | 3.683 | **<0.001** | 74.08 | 10.85-1578.96 |
| Model 2<sup>b</sup> | (Intercept) | 2.060 | 0.879 | 2.343 | 0.019 | 7.84 | 1.48-48.70 |
|  | Age | 0.026 | 0.022 | 1.195 | 0.232 | 1.03 | 0.99-1.08 |
|  | Gender male | 0.509 | 0.620 | 0.821 | 0.412 | 1.66 | 0.50-5.87 |
|  | Onset >5 | -1.525 | 0.598 | -2.552 | **0.011** | 0.22 | 0.06-0.69 |
|  | Dysgeusia | -1.436 | 0.611 | -2.350 | **0.019** | 0.24 | 0.07-0.77 |
|  | Dyspnea | -1.367 | 0.740 | -1.848 | **0.065** | 0.26 | 0.06-1.14 |

<sup>a</sup> Model 1 for the whole study population (N = 128): dependent variable =N-gene RT-PCR positivity status in Oracol sample; explanatory variables = age (continuous variable), gender (binary), symptomatic status (binary), N-gene RT-PCR Ct-value in nasopharyngeal swab (binary, "low"/"high" using a Ct threshold of 30).
<sup>b</sup> Model 2 for symptomatic participants (N = 117): dependent variable =N-gene RT-PCR positivity status in Oracol sample; explanatory variables = age (continuous variable), gender (binary), symptom onset status (binary, sampling less or more than 5 days after beginning of symptoms), dysgeusia (binary), dyspnea (binary).

## Discussion

Since the purpose of the study was to evaluate the performance of the Oracol saliva sampling device to detect SARS-CoV-2 when the corresponding nasopharyngeal swab is positive, only 128 participants were included in the analysis out of the initial 134. A short delay might have occurred between the first test used to define enrollment in the CAMOSTAT study and the samples (nasopharyngeal swab and Oracol) taken for the Oracol study, which could explain the discrepancy for the remaining six that were found negative in their second nasopharyngeal swab.

The differences between the three RT-qPCR targets for nasopharyngeal swabs and especially the lower sensitivity of the E-gene and RdRp-gene RT-PCRs cannot be explained by the quality of the samples (results for housekeeping gene not shown) since all were performed on the same RNA extract. The different assays were validated in nasopharyngeal swabs and oral fluids spiked with a serial dilution of SARS-CoV-2 virus and both sample types showed similar limits of detection (data not shown). Differences observed for clinical samples might be due to differences in the actual copy numbers of targets present in the sample, since viral RNA molecules other than the genomic RNA might be present, such as mRNAs and subgenomic RNAs. In particular, it was shown that subgenomic RNA molecules of the SARS-CoV-2 N gene are the most abundantly produced during viral replication [12]. These results nonetheless demonstrated the importance of selecting the right target for a given sample type. While the E-gene and RdRp-gene RT-qPCR showed sufficient clinical sensitivity for nasopharyngeal swabs, they were clearly not suitable for Oracol saliva samples. However, using the three assays together or the three genes within a single assay could be envision to increase the clinical sensitivity of Oracol [13].

Our logistic regression analysis on the whole study population showed that Oracol saliva samples could be a suitable alternative to nasopharyngeal swab to identify people susceptible to be shedding infectious virus, as defined by having a N-gene RT-qPCR Ct value below 30 in the nasopharyngeal swab [14–18]. Similarly, when considering only the symptomatic patients, our results showed that oral fluid collected with Oracol might be a suitable alternative to nasopharyngeal swabs for symptomatic patients sampled less than 5 days after onset of symptoms, or for those not presenting dysgeusia or dyspnea.

Oracol saliva sampling for the detection of SARS-Cov-2 has not yet been used in many studies. Interestingly, although Defeche et al. [19] found that detection of SARS-CoV-2 in Oracol samples was lower than in nasopharyngeal swab in general, it was performing better for patients presenting with fever. On the other hand, Johnson et al. [20] did not find any association between symptom type and positivity in a saliva sample, but their study included a more limited number of participants. Manabe et al. [21] also found that Oracol sampling performs best in the first five days after symptom onset, but they also suggested that the moment of sampling in the day and the addition of sputum in the sample might be worth investigating to increase the sensitivity of oral fluid samples. Our results are difficult to compare with other studies that used other devices to collect saliva or oral fluid [22]. Nonetheless, Congrave-Wilson et al. [23] also found that saliva sample was sensitive for SARS-CoV-2 detection in symptomatic patients within the first days after symptom onset and with a high viral load as estimated by RT-qPCR in the paired nasopharyngeal swab.

Our study has some limitations. First, the performance of Oracol was evaluated only in comparison of nasopharyngeal swab. Oracol was not evaluated on its own for its clinical sensitivity to detect SARS-CoV-2 infected people [6, 24]. Then, the participants to the study were mainly symptomatic patients and the observed performance might differ for asymptomatic infections [25, 26]. In addition, the samples were taken while the alpha variant was circulating in Belgium, so before the circulation of the delta and omicron variants. The performance of Oracol would need to be evaluated for these variants, especially with recent reports suggesting that the omicron variant would be detected more easily in saliva samples than nasopharyngeal swabs [27]. Finally, samples were taken before vaccination was introduced. It remains to be checked with infected, vaccinated people whether or not vaccination affects the capacity to detect SARS-CoV-2 in an Oracol sample following, for example, reduction of the viral load during infection [28–30].

In conclusion, Oracol device represents an interesting alternative to nasopharyngeal swab to detect SARS-CoV-2-infected people at a moment when they might be contagious, and should be considered as an interesting tool in the sampling strategy implemented by the health authorities to control the pandemic, especially when nasopharyngeal swabbing is not well tolerated.

## Data Availability

All data produced in the present work are contained in the manuscript

## Acknowledgements

We would to thank study nurse team of Ghent University Hospital consisting of Sophie Degroot, Sabine Buysse, Liesbeth Delesie, Lucas Van Dooren and Sophie Vanherrewege for effective and friendly patient recruitment and sampling, as well as medical laboratory technicians from Sciensano, Mona Abady, and Ghent University Hospital, Eveline Nys, Kaat D’hooghe and Sofie Blomme, for excellent laboratory work.

## References

1. Hu B, Guo H, Zhou P, Shi ZL. Characteristics of SARS-CoV-2 and COVID-19. Nat Rev Microbiol. 2021 Mar;19(3):141–154.

2. Sheridan C. Fast, portable tests come online to curb coronavirus pandemic. Nat Biotechnol. 2020 May;38(5):515–518.

3. World Health Organization (WHO) summary of available SARS-CoV-2 in-house PCR protocols https://www.who.int/docs/default-source/coronaviruse/whoinhouseassays.pdf; status on 15th of March 2020.

4. Iwasaki S, Fujisawa S, Nakakubo S, Kamada K, Yamashita Y, Fukumoto T, et al. Comparison of SARS-CoV-2 detection in nasopharyngeal swab and saliva. J Infect. 2020 Aug;81(2):e145–e147.

5. To KK, Tsang OT, Yip CC, Chan KH, Wu TC, Chan JM, et al. Consistent Detection of 2019 Novel Coronavirus in Saliva. Clin Infect Dis. 2020 Jul 28;71(15):841–843.

6. Azzi L, Carcano G, Gianfagna F, Grossi P, Gasperina DD, Genoni A, et al. Saliva is a reliable tool to detect SARS-CoV-2. J Infect. 2020 Jul;81(1):e45–e50.

7. Williams E, Bond K, Zhang B, Putland M, Williamson DA. Saliva as a Noninvasive Specimen for Detection of SARS-CoV-2. J Clin Microbiol. 2020 Jul 23;58(8):e00776–20.

8. Wyllie AL, Fournier J, Casanovas-Massana A, Campbell M, Tokuyama M, Vijayakumar P, et al.. Saliva or Nasopharyngeal Swab Specimens for Detection of SARS-CoV-2. N Engl J Med. 2020 Sep 24;383(13):1283–1286.

9. Hutse V, Van Hecke K, De Bruyn R, Samu O, Lernout T, Muyembe JJ, Brochier B. Oral fluid for the serological and molecular diagnosis of measles. Int J Infect Dis. 2010 Nov;14(11):e991–7.

10. Vyse AJ, Cohen BJ, Ramsay ME. A comparison of oral fluid collection devices for use in the surveillance of virus diseases in children. Public Health. 2001 May;115(3):201–7.

11. Tobback E, Degroote S, Buysse S, Delesie L, Van Dooren L, Vanherrewege S, et al. Efficacy and safety of camostat mesylate in early Covid-19 disease in an ambulatory setting: A randomized placebo-controlled phase II trial. Int J Infect Dis 2022 Jul 5;S1201-9712(22)00388-5.

12. Long S. SARS-CoV-2 Subgenomic RNAs: Characterization, Utility, and Perspectives. Viruses. 2021 Sep 24;13(10):1923.

13. Sun Q, Li J, Ren H, Pastor L, Loginova Y, Madej R, et al. Saliva as a testing specimen with or without pooling for SARS-CoV-2 detection by multiplex RT-PCR test. PLoS One. 2021 Feb 23;16(2):e0243183.

14. Bruce EA, Mills MG, Sampoleo R, Perchetti GA, Huang ML, Despres HW, et al. Predicting infectivity: comparing four PCR-based assays to detect culturable SARS-CoV-2 in clinical samples. EMBO Mol Med. 2022 Feb 7;14(2):e15290.

15. Bullard J, Dust K, Funk D, Strong JE, Alexander D, Garnett L, et al. Predicting Infectious Severe Acute Respiratory Syndrome Coronavirus 2 From Diagnostic Samples. Clin Infect Dis. 2020 Dec 17;71(10):2663–2666.

16. Singanayagam A, Patel M, Charlett A, Lopez Bernal J, Saliba V, Ellis J, Ladhani S, Zambon M, Gopal R. Duration of infectiousness and correlation with RT-PCR cycle threshold values in cases of COVID-19, England, January to May 2020. Euro Surveill. 2020 Aug;25(32):2001483.

17. Cevik M, Tate M, Lloyd O, Maraolo AE, Schafers J, Ho A. SARS-CoV-2, SARS-CoV, and MERS-CoV viral load dynamics, duration of viral shedding, and infectiousness: a systematic review and meta-analysis. Lancet Microbe. 2021 Jan;2(1):e13–e22.

18. La Scola B, Le Bideau M, Andreani J, Hoang VT, Grimaldier C, Colson P, et al. Viral RNA load as determined by cell culture as a management tool for discharge of SARS-CoV-2 patients from infectious disease wards. Eur J Clin Microbiol Infect Dis. 2020 Jun;39(6):1059–1061.

19. Defêche J, Azarzar S, Mesdagh A, Dellot P, Tytgat A, Bureau F, et al. In-Depth Longitudinal Comparison of Clinical Specimens to Detect SARS-CoV-2. Diagn Microbiol Infect Dis. 2022 Mar;102(3):115616.

20. Johnson AJ, Zhou S, Hoops SL, Hillmann B, Schomaker M, Kincaid R, et al. Saliva Testing Is Accurate for Early-Stage and Presymptomatic COVID-19. Microbiol Spectr. 2021 Sep 3;9(1):e0008621.

21. Manabe YC, Reuland C, Yu T, Azamfirei R, Hardick JP, Church T, et al. Self-Collected Oral Fluid Saliva Is Insensitive Compared With Nasal-Oropharyngeal Swabs in the Detection of Severe Acute Respiratory Syndrome Coronavirus 2 in Outpatients. Open Forum Infect Dis. 2020 Dec 30;8(2):ofaa648.

22. Warsi I, Khurshid Z, Shazam H, Umer MF, Imran E, Khan MO, et al. Saliva Exhibits High Sensitivity and Specificity for the Detection of SARS-COV-2. Diseases. 2021 May 20;9(2):38.

23. Congrave-Wilson Z, Lee Y, Jumarang J, Perez S, Bender JM, Bard JD, Pannaraj PS. Change in Saliva RT-PCR Sensitivity Over the Course of SARS-CoV-2 Infection. JAMA. 2021 Sep 21;326(11):1065–1067.

24. Jamal AJ, Mozafarihashjin M, Coomes E, Powis J, Li AX, Paterson A, et al. Sensitivity of Nasopharyngeal Swabs and Saliva for the Detection of Severe Acute Respiratory Syndrome Coronavirus 2. Clin Infect Dis. 2021 Mar 15;72(6):1064–1066.

25. Teo AKJ, Choudhury Y, Tan IB, Cher CY, Chew SH, Wan ZY, et al. Saliva is more sensitive than nasopharyngeal or nasal swabs for diagnosis of asymptomatic and mild COVID-19 infection. Sci Rep. 2021 Feb 4;11(1):3134.

26. Tutuncu EE, Ozgur D, Karamese M. Saliva samples for detection of SARS-CoV-2 in mildly symptomatic and asymptomatic patients. J Med Virol. 2021 May;93(5):2932–2937.

27. Marais G, Hsiao NY, Iranzadeh A, Doolabh D, Enoch A, Chu CY, et al, Saliva swabs are the preferred sample for Omicron detection. MedRxiv, December 24, 2021.

28. Levine-Tiefenbrun M, Yelin I, Alapi H, Katz R, Herzel E, Kuint J, Chodick G, Gazit S, Patalon T, Kishony R. Viral loads of Delta-variant SARS-CoV-2 breakthrough infections after vaccination and booster with BNT162b2. Nat Med. 2021 Dec;27(12):2108–2110.

29. Puhach O, Adea K, Hulo N, Sattonnet P, Genecand C, Iten A, al. Infectious viral load in unvaccinated and vaccinated patients infected with SARS-CoV-2 WT, Delta and Omicron. MedRxiv, January 18, 2022.

30. Acharya CB, Schrom J, Mitchell AM, Coil DA, Marquez C, Rojas S, et al. No Significant Difference in Viral Load Between Vaccinated and Unvaccinated, Asymptomatic and Symptomatic Groups When Infected with SARS-CoV-2 Delta Variant. MedRxiv, October 5, 2021.

